# Keynote 48: Is it really for everyone?

**DOI:** 10.1101/2020.04.18.20070888

**Authors:** Jonathan D. Schoenfeld, Geoffrey Fell, Robert I. Haddad, Lorenzo Trippa

**Affiliations:** Brigham and Women’s Hospital; Dana-Farber Cancer Institute and Harvard Medical School, Boston, MA

## Abstract

Burtness et al. recently published the landmark Keynote-48 study, demonstrating a survival benefit for pembrolizumab monotherapy and pembrolizumab/chemotherapy, compared with cetuximab/chemotherapy, in patients with recurrent/metastatic head and neck squamous cell carcinoma (HNSCC). These data are impactful and practice-changing, and have rapidly been adopted in practice, with increasing numbers of HNSCC patients receiving either pembrolizumab monotherapy, or pembrolizumab / chemotherapy, in the first line recurrent / metastatic setting. Pembrolizumab was approved as a single agent for patients whose tumors express PD□L1 (Combined Positive Score [CPS] ≥1), while pembrolizumab / chemotherapy was approved for use in the United States for all patients irrespective of PD-L1 expression.

---

Burtness et al. recently published the landmark Keynote-48 study, demonstrating a survival benefit for pembrolizumab monotherapy and pembrolizumab/chemotherapy, compared with cetuximab/chemotherapy, in patients with recurrent/metastatic head and neck squamous cell carcinoma (HNSCC).^1^ These data are impactful and practice-changing,^2^ and have rapidly been adopted in practice, with increasing numbers of HNSCC patients receiving either pembrolizumab monotherapy, or pembrolizumab / chemotherapy, in the first line recurrent / metastatic setting. Pembrolizumab was approved as a single agent for patients whose tumors express PD□L1 (Combined Positive Score [CPS] ≥1), while pembrolizumab / chemotherapy was approved for use in the United States for all patients irrespective of PD-L1 expression.^3^

In the Keynote-48 study, based on an amended design, overall survival comparisons were made in the PD-L1 CPS >=20, >=1, and total populations; these results defined subsequent regulatory approval. However, these results offer little specific information describing outcomes specifically in patients with low CPS, the suitability of the CPS thresholds chosen, and hypotheses to be tested in future clinical trials.

To address these gaps, we extracted data from Kaplan-Meier curves^4^ and estimated survival in the CPS<1 population (Figure), which was not reported in the manuscript. This was done by leveraging the fact that the overall survival function is a weighted average of CPS < and >= 1. These data suggest cetuximab/chemotherapy may retain benefit in PD-L1 negative tumors, an observation of potential clinical relevance directly affecting patient care. This approach also illustrates the possibility, in some cases, of estimating group-specific survival distributions for subpopulations of interest even when Kaplan-Meier curves are not provided that can be applied to other trials.

**Figure (below):**
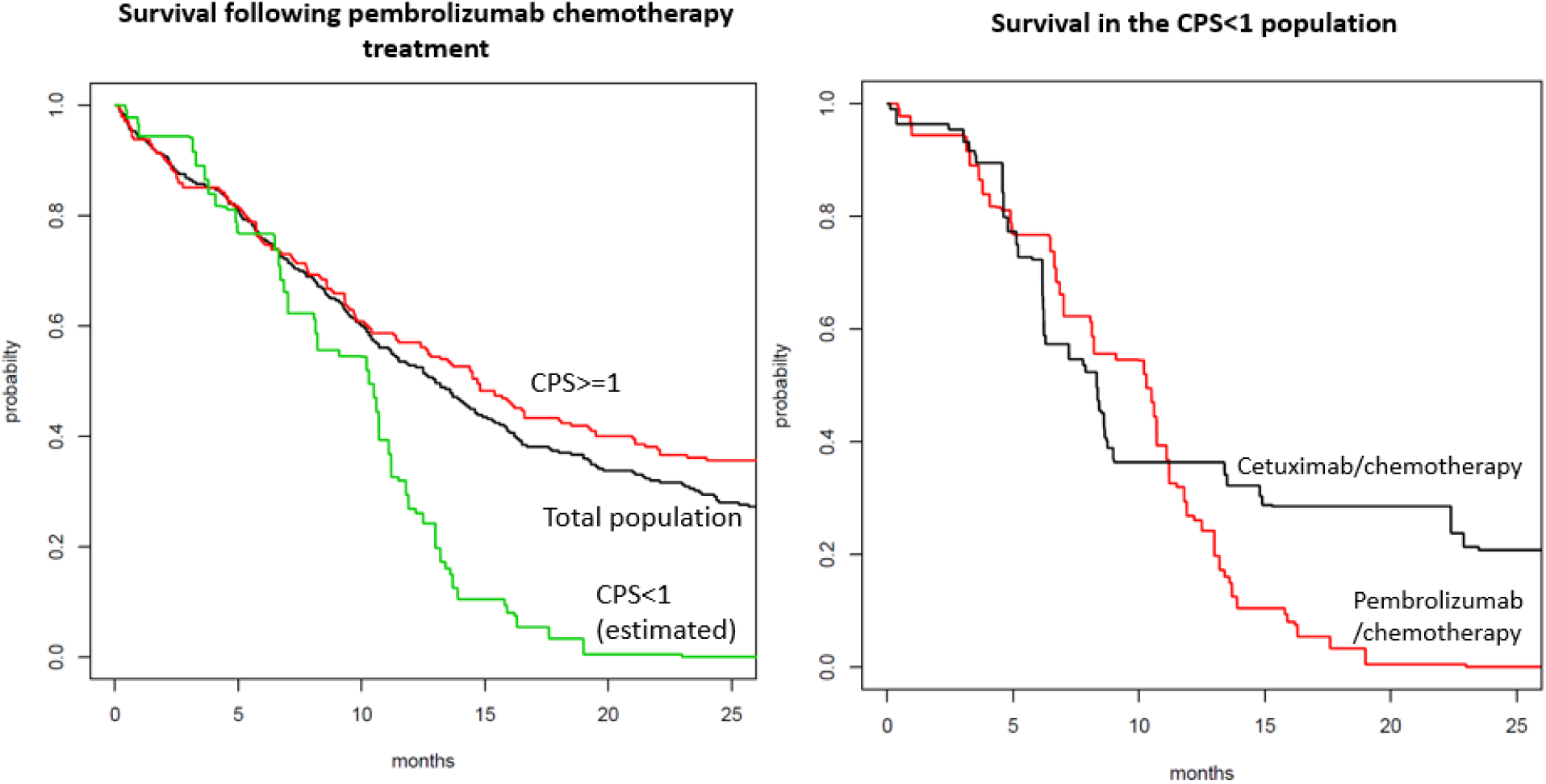
<u>Left:</u> Using an approach outlined in [4], we extracted Kaplan-Meier curves from the pembrolizumab/chemotherapy population and used the survival.function-CPS>=1.0 (red) survival.function-overall.population (black) to estimate survival.function-CPS<1.0 (green, *not reported in the original manuscript*) using the equation: weight × survival.function-CPS<1.0 + (1-weight) × survival.function-CPS>=1.0 =survival.function.overall.pop In this case with weight 39/281. <u>Right:</u> We compare overall survival following treatment with pembrolizumab/chemotherapy (red) to cetuximab/chemotherapy (black) in the CPS<1 population. Survival functions were estimated as in the left panel.

## Data Availability

Not applicable

